# Climate variation and serotype competition drive dengue outbreak dynamics in Singapore

**DOI:** 10.1101/2024.09.17.24313793

**Authors:** Emilie Finch, Adam Kucharski, Shuzhen Sim, Lee-Ching Ng, Rachel Lowe

## Abstract

Dengue poses a rapidly increasing threat to global health, with Southeast Asia as one of the worst affected regions. Climate-informed early warning systems can help to mitigate the impact of outbreaks; however, prediction of large outbreaks with sufficient lead time to guide interventions remains a challenge. In this work, we quantify the role of climatic variation and serotype competition in shaping dengue risk in Singapore using over 20 years of weekly case data. We integrated these findings into an early warning system framework able to predict dengue outbreaks up to 2 months ahead. While a climate-informed model improved predictive power by 54% compared to a seasonal baseline, including additional serotype information increased predictive performance to 60%, helping to explain interannual variation. By incorporating serotype competition as a proxy for population immunity, this work advances the field of dengue prediction and demonstrates the value of long-term virus surveillance.

## Introduction

Climate-informed forecasting models can be used to give advance warning of infectious disease outbreaks and mitigate their impact. Early warning systems integrating climate information are becoming an increasingly important tool for epidemic response as the climate crisis leads to more frequent climatic extremes and shifts the dynamics of climate-sensitive infectious diseases, such as dengue (1,2).

Dengue is an emerging vector-borne disease, transmitted by *Aedes aegypti* and *Aedes albopictus* in urban and peri-urban areas (3). Global reported dengue incidence has increased by 30-fold over the past 50 years, alongside increases in the geographical range of transmission, and approximately half of the global population is thought to be at risk of dengue transmission (4). There are four antigenically distinct serotypes of dengue virus (DENV1-4): infection with one serotype results in life-long immunity to the infecting serotype, and limited cross-immunity to others (5,6). Dengue epidemic dynamics are complex and challenging to predict, with large outbreaks driven by multiple factors including climate variation, competition between the four dengue serotypes and traditional and novel vector-control efforts.

Singapore, an equatorial city-state in Southeast Asia, experiences hyperendemic dengue transmission with all four serotypes in circulation and cyclical replacements in the dominant serotype (7). Singapore experiences warm and humid temperatures year round, with suitable conditions for mosquito breeding and dengue transmission. Rainfall is affected by two monsoon seasons, with the Northeast monsoon occurring from December until early March and the Southwest monsoon occurring from June to September (8). The peak dengue season usually occurs between June and October, typically following the warmest and most humid months of the year. Since the 1960s, Singapore has implemented stringent dengue prevention measures focused on vector control and public education. This has led to a reduction in the *Aedes* House Index (AHI, a measure of the percentage of houses positive for *Aedes* breeding) from 48% in 1966 to around 1% in the 1990s (9). Periodic seroprevalence surveys have demonstrated a concurrent decrease in seroprevalence in almost all age groups, with an estimated decrease in force of infection (FOI) from around 0.1 per year in the 1960s to 0.01 per year from the 1990s onwards (10,11). This is also reflected in surveillance data showing an increase in the average age of reported cases from children to young adults (7,9,12). Despite this, reported cases have increased in recent years. Possible explanations for this include improved case detection and reporting, particularly after 2008 where a campaign was launched encouraging the use of NS1 rapid tests in laboratories, or higher population vulnerability to dengue outbreaks (for example, after the importation of a new viral genotype) due to lower immunity (9,11).

Singapore experiences cyclical dengue outbreaks which have been increasing in frequency and magnitude (7,9). These have followed switches in dominant circulating serotypes, with the exception of the 2019 outbreak. Between 2006 and 2020, virus surveillance showed the dominant serotype switching between DENV1 and DENV2 (7,13). Accordingly, population immunity to DENV1 and DENV2 is believed to be higher than against DENV3 and DENV4. For example, a study of healthy blood donors in 2009 amongst 16 to 60 year olds found seropositivity of 35.8% anti-DENV1 and 36.4% anti-DENV2 compared with 15.4% anti-DENV3 and 7.7% anti-DENV4 (9,10). However, the cyclical pattern of DENV1 and 2 dominance has been disrupted in recent years. A large, predominantly DENV2 outbreak in 2019-2020 saw an increasing contribution from DENV3, which then gained predominance in the next outbreak year in 2022 (14). Dengue outbreaks have also been associated with El Niño events, the warm phase of the El Niño Southern Oscillation (ENSO), involving warmer than normal oceanic and atmospheric temperatures in the Pacific, which typically lead to hotter and drier climatic conditions in Singapore (15–17).

Climate influences dengue transmission through effects on the vector (*Aedes* mosquitos) and the dengue virus itself. Temperature affects mosquito survival, development and reproduction, as well as the viral extrinsic incubation period, with an optimal temperature for transmission of around 29°C and thermal limits between 17.8°C – 34.5°C (18). Temperature has been found to shape the timing, length and geographical extent of dengue seasons (2,19). Contrastingly, the impact of rainfall on dengue transmission is more nuanced. While increasingly wet and humid conditions can lead to the creation of mosquito breeding sites, the effect of rainfall on transmission is dependent on human water storage behaviour, and the availability of water and sanitation infrastructure, which can lead to non-linear and delayed impacts of rainfall on dengue transmission (20–22). In particular, excessive rainfall can lead to flushing effects, where mosquito breeding habitats are washed away entirely. This has been documented in Singapore, where dengue outbreak risk was found to decrease following flushing events (23). Previous research in Singapore has demonstrated the utility of temperature, precipitation and absolute humidity in predicting dengue incidence (24–26). Currently, Singapore uses a machine learning approach based on LASSO regression to generate operational forecasts of dengue incidence for outbreak alerts and decision-support (27).

Forecasting models can capitalise on inherent lags between climatic variation and dengue transmission to predict outbreak risk at operationally useful lead times (28). In Singapore, an analysis of vector control found that local authorities needed an average of 2 months to mitigate the impact of a dengue outbreak, suggesting that early warning forecasts with several months lead time would be optimal (29). Forecasts at shorter horizons may also be helpful to inform situational awareness. To date, dengue forecasting models have struggled to predict interannual variability in dengue seasons and shown worse predictive performance for high incidence seasons, which have the greatest public health impact (30). Additionally, forecast skill is typically lower earlier in the season when aiming to forecast several months ahead, which is when forecasts have the most potential operational value. While immunity is theoretically recognised as an important driver of interannual dengue dynamics, to our knowledge no current dengue forecasting models directly account for serotype dynamics or changes in immunity, which are likely to be particularly important in hyperendemic regions such as Southeast Asia. To address this, we incorporate climate and serotype dynamics within a Bayesian hierarchical modelling framework to forecast dengue incidence. We quantify the effect of climatic variables, the Niño 3.4 index and switches in dominant serotype on dengue incidence, and evaluated forecasts of dengue incidence with a 2-8 week forecast horizon.

## Results

### Reported dengue cases in Singapore over the past two decades

Between 1 January 2000 and 31 December 2022, 234,358 cases of dengue were reported in Singapore. Most cases were reported between June and September, typically following warm and humid climatic conditions (Figure 1, panels D-F). Cyclical outbreaks, which historically were thought to occur every 5-6 years, have become more frequent and of larger magnitude (Figure 1). For instance, while the 2004 dengue outbreak led to 9,459 cases overall and peaked at 332 reported weekly cases, the 2022 outbreak resulted in 32,259 reported cases and peaked at 1,568 reported weekly cases. We defined an outbreak week using a seasonal moving 75th percentile threshold. For a given month and year, we defined an outbreak threshold of the 75th percentile of weekly cases in that month using all years up to (but not including) the given year. This identifies the same outbreak periods as the endemic channel threshold used within the NEA but has several added benefits. Firstly, a percentile threshold is simpler to calculate and adjusts on a rolling basis rather than year-on-year. Additionally, by incorporating the seasonal patterns underlying dengue transmission, this definition allows outbreak periods to be defined earlier than with a fixed, non-seasonal threshold (Supplementary information, Figure 1).

**Figure 1:**
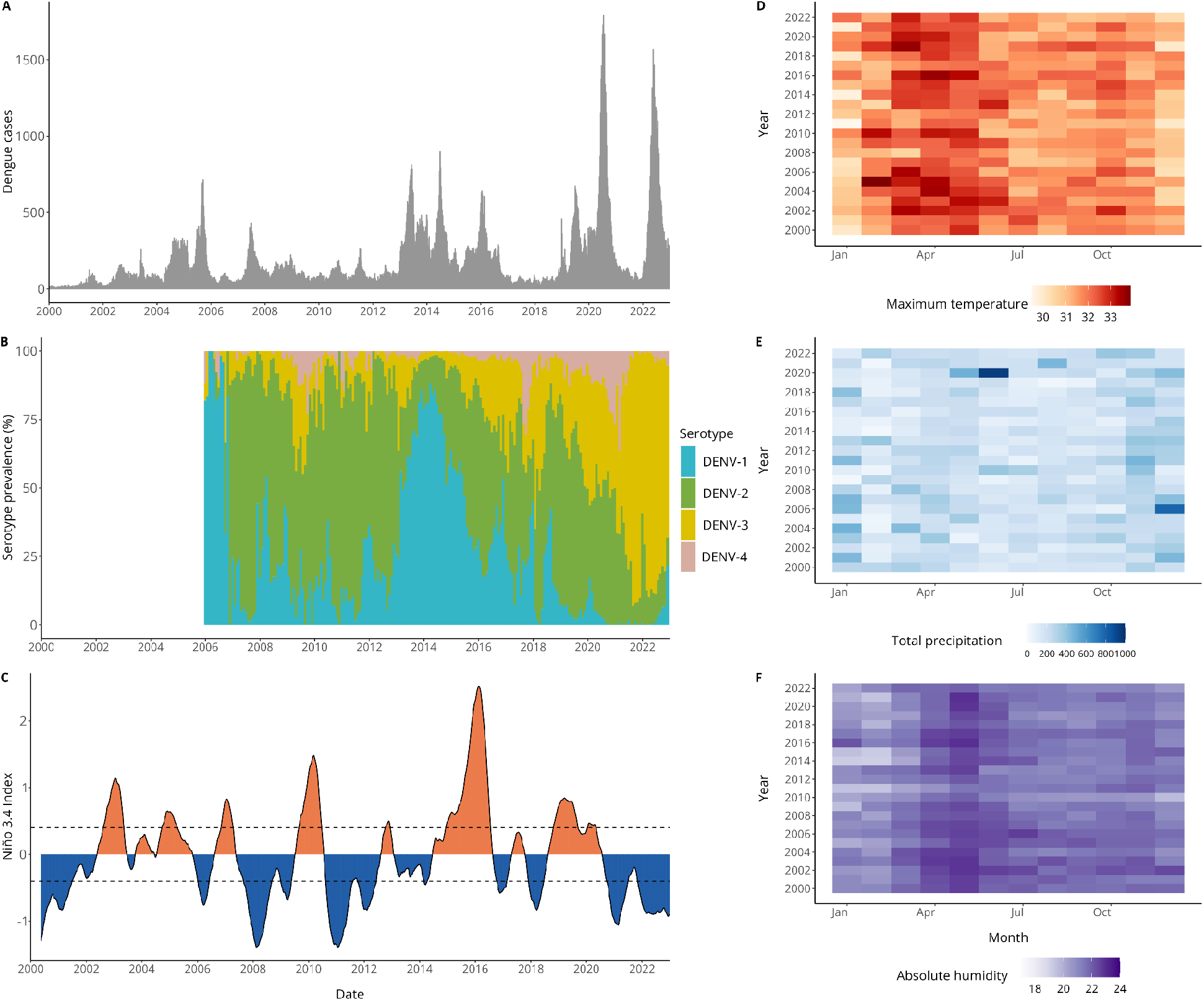
Dengue cases, climate variability and serotype dominance in Singapore. Figure showing epidemiological and climatic data for Singapore from 2000 – 2023. A) Bars show weekly reported dengue cases from 1 January 2000 – 31 December 2022. B) Stacked bars show the proportion of each serotype detected through virus surveillance. To calculate this, we aggregate the number of serotyped cases for DENV1-4 at a monthly level and divide each by the total number of cases serotyped in that month. C) Line graph showing weekly Niño 3.4 sea surface temperature anomalies, dashed lines indicate +0.5°C and -0.5°C which are often used as thresholds to define El Niño and La Niña events. Blue shading indicates negative values of the Niño 3.4 index (indicating La Niña conditions), while orange shading indicates positive values, associated with El Niño conditions. Heatmaps for (D) monthly mean maximum temperature (°C), (E) total precipitation (mm), and (F) mean absolute humidity (g/m^3^).

We then defined an outbreak year as a year containing more than 12 outbreak weeks. This identified outbreaks in; 2004, 2005, 2007, 2013, 2014, 2015, 2016, 2019, 2020 and 2022. In some years, dengue outbreaks coincided with switches in dominant serotype; such as the switch from DENV-2 to DENV-1 in 2013 or from DENV-2 to DENV-3 in 2022 (Figure 1). Additionally, outbreaks sometimes coincided with El Niño events, defined when sea surface temperature (SSTs) in the Niño 3.4 region exceed 0.5°C for 6 consecutive months.

### Climate and serotype dynamics shape dengue risk with non-linear and delayed effects

To quantify the effect of climate, ENSO, and changes in serotype on dengue risk, we fit a Bayesian hierarchical model to weekly case data. We used a negative binomial likelihood and incorporated weekly random effects to capture seasonality and yearly random effects to account for unexplained interannual variation in dengue risk, for instance, due to control measures (Methods). We then compared a baseline model (including only weekly and yearly random effects) with models containing climatic and serotype covariates to see whether their inclusion improved model adequacy statistics, and reduced unexplained interannual variation in the model. We tested temperature (non-linear and linear), precipitation (non-linear and linear), humidity (linear) and ENSO (non-linear and linear) variables considering lags from 0:20 weeks, as well as serotype variables (non-linear and linear). We selected a final model including: 12-week rolling average maximum temperature in °C; 12-week total days without rain; 12-week rolling average Niño 3.4 SSTA with a 4 week lag; and a time-varying covariate measuring the number of weeks since a switch in dominant serotype (Methods). We found a non-linear relationship between maximum temperature and dengue incidence risk, with increased risk around 32 °C and decreased risk at particularly low or high maximum temperatures (Figure 2, panel A). Similarly, we found increased risk of dengue at intermediately wet conditions, with around 30 days without rain in the previous 3 months, and decreased risk in dry conditions, with more than 45 days without rain in the previous 3 months (Figure 2, panel B). We found decreased dengue risk with negative Niño 3.4 SSTA values and non-linearly increasing dengue risk with values of Niño 3.4 SSTA upwards of around 1.4 (Figure 2, panel C). Finally, we found a non-linear relationship between the time since a switch in dominant serotype and dengue risk, with increased risk in the first two years following a switch, decreased risk between 2-6 years following a switch, and subsequent increased risk at 6+ years following a switch (Figure 2, panel D).

**Figure 2:**
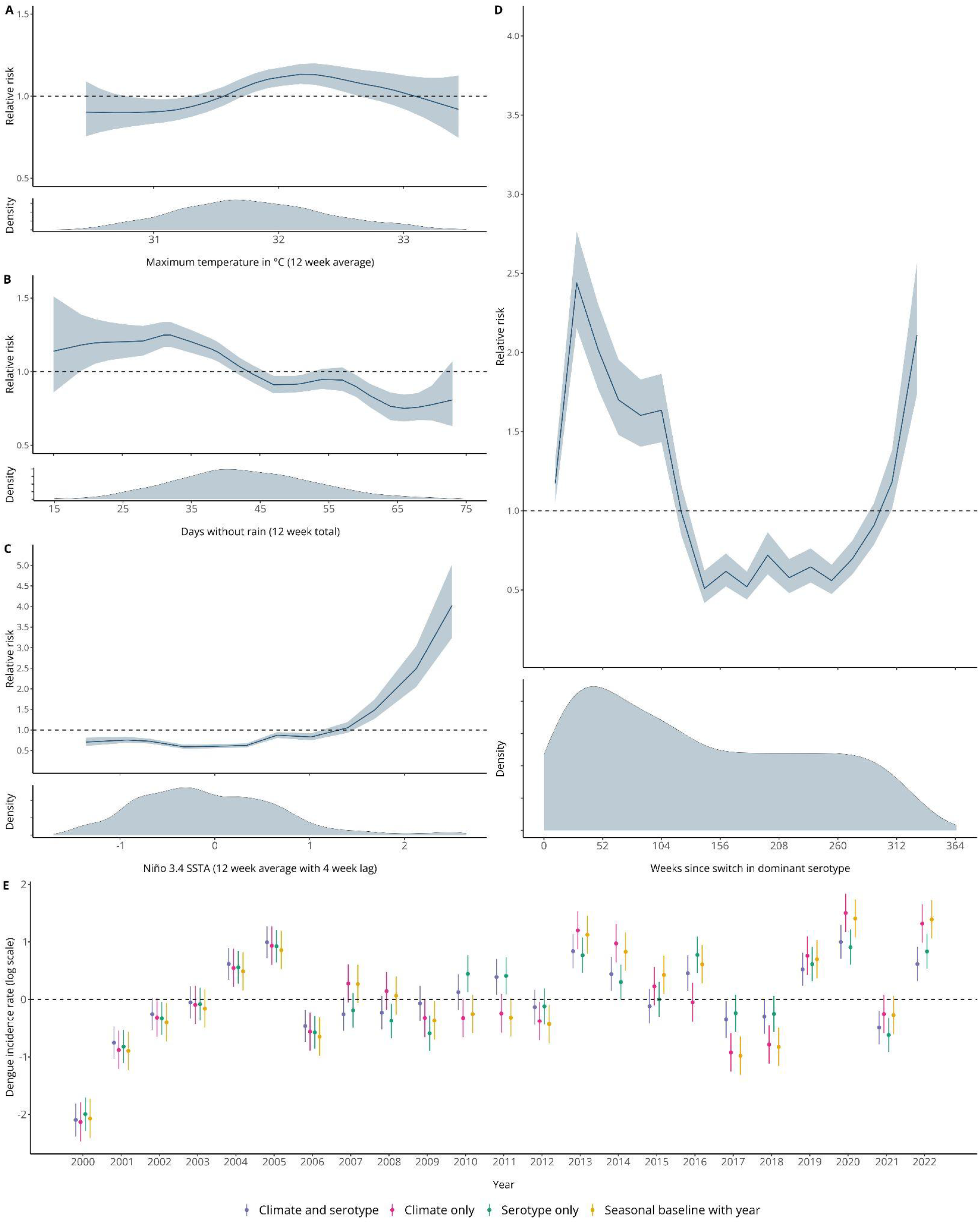
Effects of climatic variability and switches in dominant serotype on dengue incidence in Singapore. Panels A-D show posterior marginal effects and density plots for covariates in the final selected model. These include maximum temperature in °C (12 week running average), days without rain (12 week total), Niño 3.4 SSTA (12 week average with a 4 week lag) and weeks since switch in dominant serotype. These are shown on the relative risk scale displaying the median value and associated 95% credible interval and can be interpreted as the effect of the covariate on dengue incidence rate with all other parameters held constant. Panel E compares the estimated yearly random effect, γ_*a*[*t*]_, for a s*easonal baseline with year* model including only weekly and yearly random effects γ_*a*[*t*]_ + δ_*w*[*t*]_ (in yellow), the final selected c*limate and serotype* model including all climate and serotype covariates and random effects (in purple), a *climate only model* with random effects (in pink), and a *serotype only* model with random effects (in green). The estimated yearly random effect from the baseline model indicates whether dengue incidence was higher or lower for that year than the overall mean incidence. We would expect the estimated yearly random effects for covariate models to be closer to 0 (indicated with a dashed line) when covariates are able to account for interannual variability in dengue incidence.

We then compared the yearly random effects estimated for our final *climate and serotype* model, a *climate only* model including climatic covariates and random effects, a *serotype only* model including the serotype covariate and random effects, and a baseline model including weekly and yearly random effects (Figure 2, panel E). As yearly random effects account for unexplained interannual variation in dengue incidence, we would expect these values to be closer to 0 when other model covariates are able to explain this variation. We found no difference in estimated yearly random effects between the four models before 2006, which we would expect as no serotype information is available before this date. From 2007 onwards, overall, models including serotype information tended to have yearly random effects closer to 0. By calculating the percentage reduction in mean absolute yearly random effects between covariate models and the baseline model, we can quantify how much model covariates explain interannual variation in dengue incidence (Methods). While a *climate only* model reduced unexplained interannual variation in dengue incidence by 4.1%, including additional serotype information (in the *climate and serotype* model) resulted in a 26.8% reduction. Contrastingly, a *serotype only* model reduced unexplained interannual variation by 19.4%.

### Accounting for serotype and climate dynamics improves probabilistic predictions of dengue case incidence and outbreak detection

We used a time series cross-validation approach to produce probabilistic dengue predictions and calculate the probability of exceeding a predefined outbreak threshold in a given week (31). This is an appropriate design to assess model utility for forecasting, as we preserve the underlying time-order of the data. We first generated probabilistic predictions using our candidate models with no lead time (using information up to a target date to predict dengue cases on that date) to compare out-of-sample predictive ability between 2009 and 2022 (Figure 3). The first 8 years of data were used exclusively for training (Methods). We compared our final selected *climate and serotype* model with a *climate only* model and a *serotype only* model containing only climatic and serotype covariates respectively (Supplementary information, Table 3). We compared these to a *seasonal baseline* model which only included weekly random effects. This is equivalent to a climatological baseline model which uses the average seasonal pattern in dengue incidence to predict cases in a given target week. We assessed forecast skill in predicting weekly dengue cases using the continuous ranked probability score (CRPS), where smaller values indicate better performance. We also calculated the continuous ranked probability skill score (CRPSS) which is defined as the percentage improvement in CRPS compared to a baseline model. We also assessed the predictive ability of the candidate models for outbreak detection using the Brier score and conducted receiver operating characteristic (ROC) analysis. Here, we compared model hit rate (proportion of outbreak weeks correctly identified) with false alarm rate (the proportion of weeks without an outbreak where an outbreak was predicted to occur). We calculated the area under the curve (AUC) to measure model skill in classifying outbreak and non-outbreak weeks (Methods).

**Figure 3:**
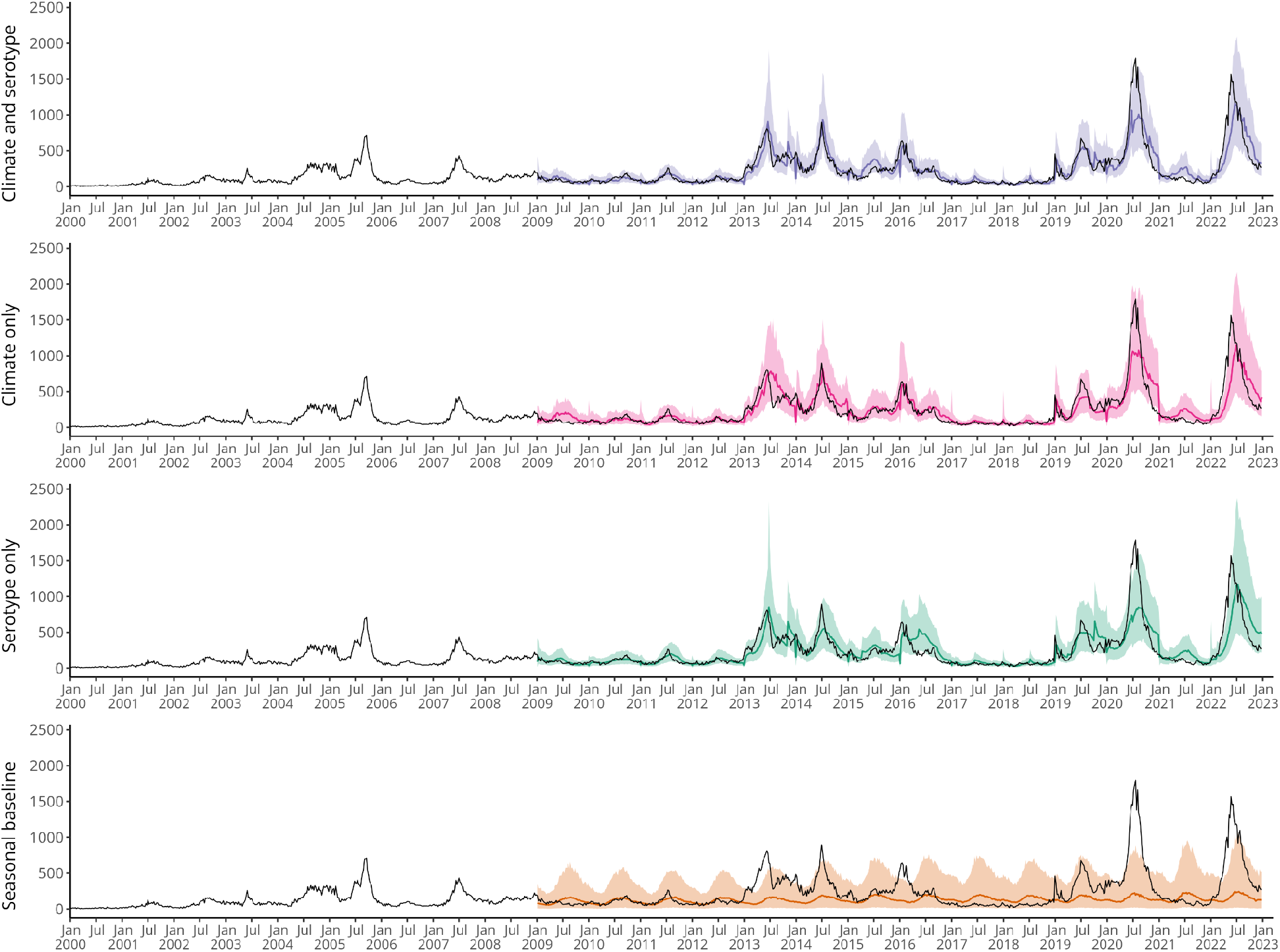
Comparing time series cross-validated predictions of candidate models. Figure showing time series cross-validated posterior predictions of dengue cases for each model from 2009 – 2022. We used an expanding window cross-validation methodology, where the model is trained on data up to but not including the target week and then posterior predictions are generated for the target week. Coloured lines show the median posterior prediction of weekly dengue cases, shaded areas show the 95% credible interval and the dark grey lines show the data. From top to bottom the figure shows: predictions for the final selected ‘*Climate and serotype*’ model with weekly and yearly random effects γ_*a*[*t*]_ + δ_*w*[*t*]_ in purple; predictions for a ‘*Climate only*’ model with weekly and yearly random effects in pink; predictions for a ‘*Serotype only*’ model with weekly and yearly random effects in green; and predictions from a ‘*Seasonal baseline*’ model with only weekly random effects δ_*w*[*t*]_in orange.

The *climate and serotype* model was able to reproduce dengue epidemic dynamics in Singapore between 2009 and 2022 and had a lower CRPS than all other models, indicating better performance. In particular, the *climate and serotype* model is better able to predict the decrease in early 2016 than a *serotype-only* model (which predicts a late peak around July 2016). Similarly, the *climate and serotype* model outperforms the *climate-only* model in predicting the decrease in cases following peaks in mid-2013 and mid-2014, as well as better predicting peak cases in July 2019. All covariate models underpredicted the peak in 2020 but were able to accurately predict peak timing. Similarly in 2022, covariate models underpredicted the peak and predicted later peak timing than what was seen. The *climate and serotype* model showed a 60% relative improvement over the *seasonal baseline* model according to the CRPSS. The *climate only* and *serotype only* models also performed well, with a 54% and 49% relative improvement over the *seasonal baseline* model respectively (Supplementary information, Table 3). The *climate and serotype* model also outperformed other models in outbreak detection, with a lower Brier score (indicating better performance). This can be seen in Figure 4, where the *climate and serotype* model is better able to assign high probability of an outbreak to actual outbreak weeks and lower probability of an outbreak to non-outbreak weeks. The *climate and serotype* model also had the highest area-under-the-curve (AUC) (98%, 95% CI: 97.7 - 99.0%) and corresponding lowest false alarm rate (2.1%) and the highest hit rate (92%) of the candidate models, with an optimal model outbreak alert threshold of 71% (Supplementary information, Table 4). The *climate only* and *serotype only* models also performed well in outbreak detection, as can be seen from the overlapping ROC curves in Figure 4.

**Figure 4:**
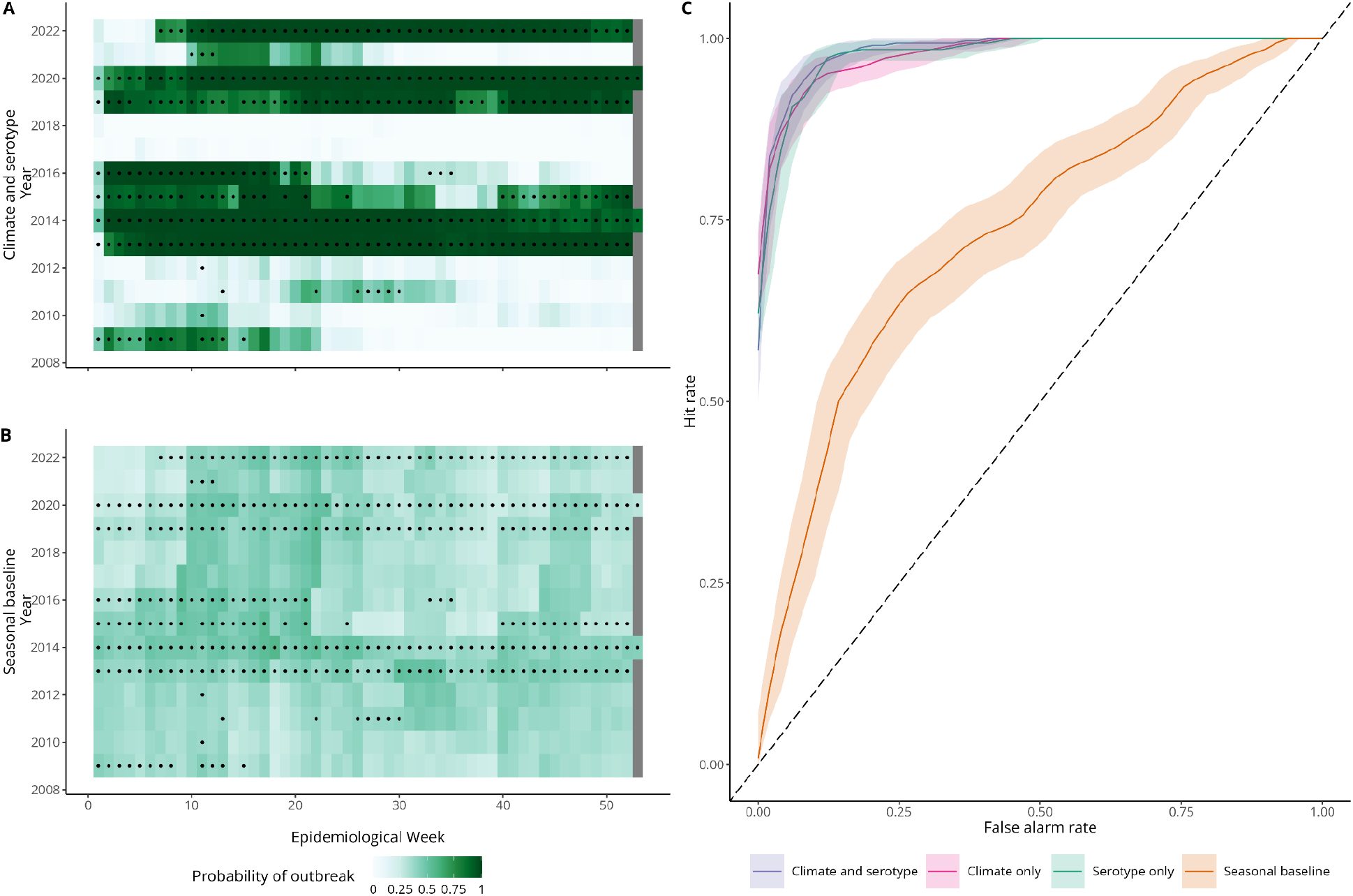
Comparing outbreak detection of candidate models. Panels A and B show tile plots of model posterior predictions of exceeding the outbreak threshold for each week between 2009 – 2022. White indicates *P*_*outbreak*_ = 0, while dark green indicates *P*_*outbreak*_ = 1. Circles indicate observed outbreak weeks defined using a seasonal moving 75th percentile threshold. For a given month and year, we defined an outbreak threshold using the 75th percentile of weekly cases in that month using all years up to, but not including, the given year. Panel E shows an ROC curve, plotting hit rate against false alarm rate for different model outbreak alert thresholds. Hit rate (or sensitivity) is defined as the proportion of outbreak weeks that were correctly predicted. False alarm rate (1 - specificity) is defined as the proportion of weeks without an outbreak where an outbreak was predicted to occur. The shaded area shows the 95% confidence interval around the ROC curve.

### Dengue forecasting for early warning with 2-8 weeks lead time

Having identified a model able to predict dengue cases and outbreak weeks with no lead time, we then adapted our framework for use in an early-warning context, producing forecasts with 2 to 8 weeks lead time. For each forecast horizon, we used the best approximation of the covariates in the final models available at the lead time considered (Methods, Supplementary information, Figure 3). We then produced probabilistic predictions of dengue using our candidate models to compare predictive ability with 2-8 week forecast horizons. Forecasts for 4 weeks ahead and 8 weeks ahead are shown in Figure 5, with forecasts for all horizons in Supplementary information, Figure 2. As expected, predictive ability declined with forecast horizon, with better performance at shorter lead times and increased uncertainty around model predictions at longer lead times (Figure 6). For instance, at an 8 week forecast horizon, the *climate and serotype* model struggled to predict peaks in late 2015-2016, first over and then under-predicting.

**Figure 5:**
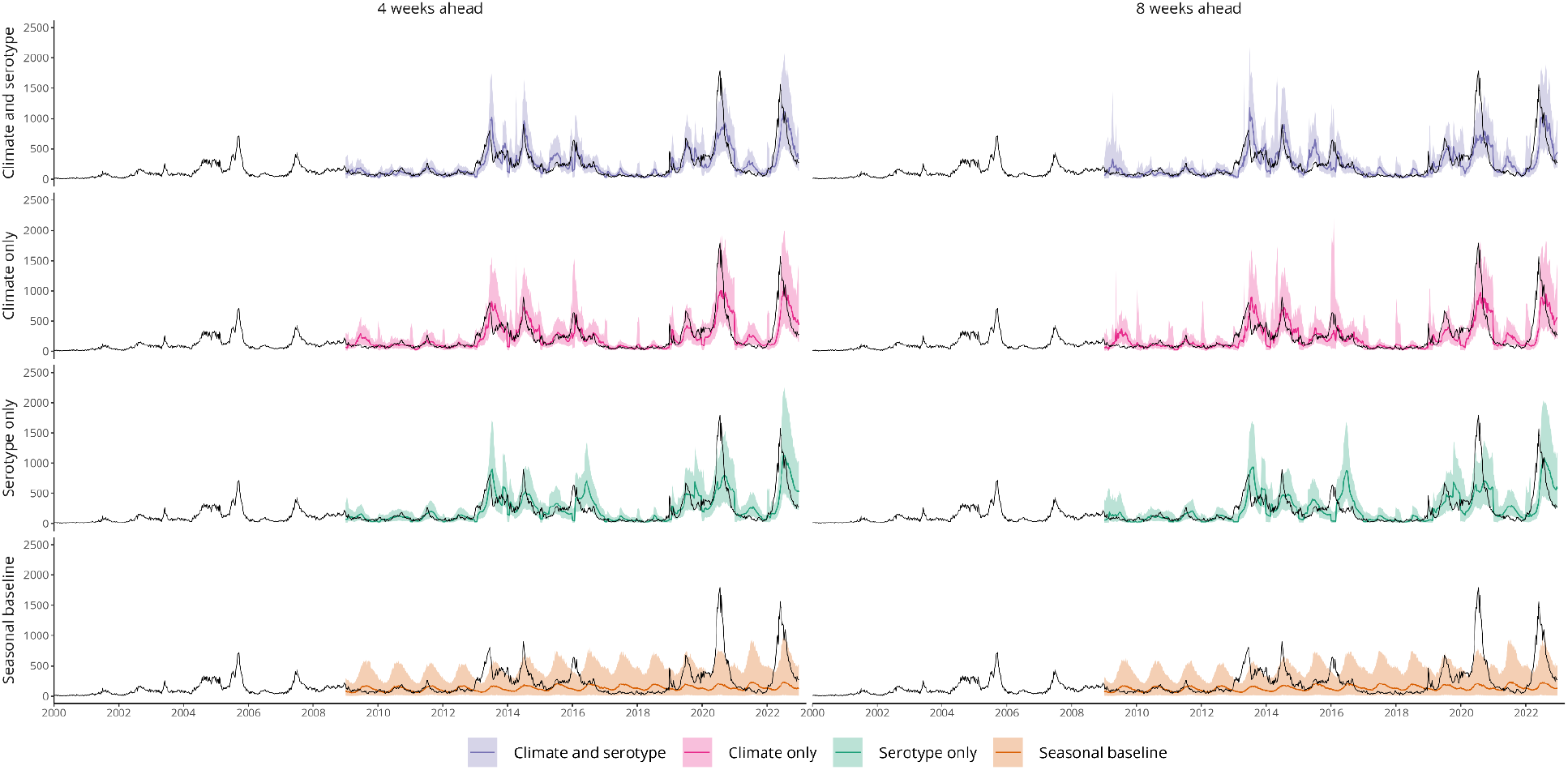
Dengue forecasts for early warning at 4 and 8 week forecast horizons. Figure showing time series cross-validated posterior predictions of dengue cases for each model from 2009 – 2022 at 2 - 8 week forecast horizons. We used an expanding window time series cross-validation methodology, where posterior predictions for each week are generated from a model fit to data up to week *t - h*, where *t* is the target week and *h* is the forecast horizon. Coloured lines show the median posterior prediction of weekly dengue cases, shaded areas show the 95% credible interval and the dark grey line shows the data. From top to bottom the figure shows: predictions for the final selected *climate and serotype* model with weekly and yearly random effects γ_*a*[*t*]_ + δ_*w*[*t*]_ in purple; predictions for a *climate only* model with weekly and yearly random effects in pink; predictions for a *serotype only* model with weekly and yearly random effects in green; and predictions from a s*easonal baseline* model with only weekly random effects δ_*w*[*t*]_ in orange. From left to right each column shows forecasts at 4 and 8 weeks ahead respectively.

**Figure 6:**
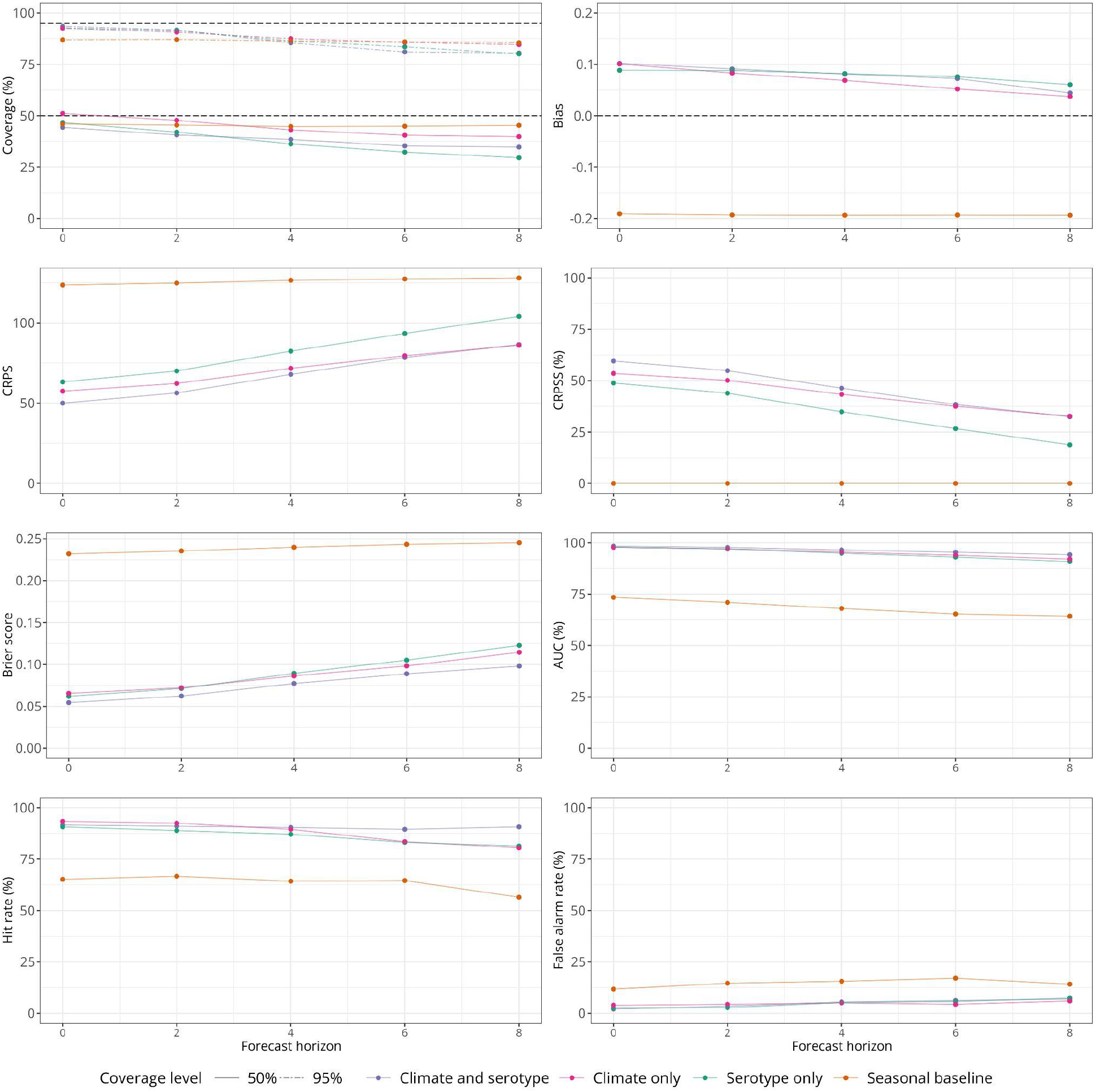
Predictive performance over different forecast horizons. Figure showing forecast metrics for each model across all forecast horizons from 2009 – 2022. From top left to bottom right these show: interval coverage %, bias; CRPS (continuous ranked probability score), CRPSS (continuous ranked probability skill score, %), Brier score, AUC (area under the curve, %), hit rate (%) and false alarm rate (%). Interval coverage shows the percentage of observations falling inside a given prediction interval. A perfectly calibrated forecast would have coverage equal to the nominal prediction interval; that is, 95% coverage equal to 95% and 50% coverage equal to 50%, indicated by dashed horizontal lines. Bias measures the relative tendency of the model to over- or under-predict, and is bounded between -1 and 1, with 0 indicating unbiased forecasts. The CRPS can take values between 0 and infinity, with smaller values indicating better performance. The CRPSS indicates the relative improvement of each covariate model over the *seasonal baseline* model and can take values from 0%, indicating that the model performs the same as the baseline, and 100%, indicating perfect forecasting skill. The Brier score can take values from 0 – 1, with smaller values indicating better performance. The AUC can take values from 0-100% with 100% indicating perfect classification. Hit rate and false alarm rate also take values from 0 – 100% with higher and lower values indicating better performance respectively.

However, all three covariate models considered were able to capture broad epidemic dynamics from 2009-2022 even at longer lead times and offered considerable improvement in performance compared with the *seasonal baseline*. For instance, at an 8 week forecast horizon the *climate and serotype* model showed 32% relative skill improvement over the *seasonal baseline.* Additionally, the *climate and serotype* model continued to accurately detect outbreak weeks at all forecast horizons, with an AUC of 94% (95% CI: 92.7 - 95.7%) at an 8 week forecast horizon (Supplementary information, Table 4).

We compared forecast skill metrics for each model from 0-8 weeks ahead and found the *climate and serotype* model had a lower CRPS than other models (indicating better performance) until a 6 week forecast horizon, when its performance was equivalent to the *climate only* model. However, when considering outbreak detection metrics (Brier score and AUC) the *climate and serotype* model outperformed all other models at all forecast horizons (Figure 6).

## Discussion

Climate and serotype dynamics have important impacts on dengue transmission and outbreak risk; however, to date few statistical forecasting models account for both drivers within the same framework. This could lead to misattribution of dengue risk caused by changes in population immunity to climatic or other non-climatic factors, and limit the ability to forecast large outbreaks in advance. Additionally, mechanistic approaches aiming to better capture the biological processes underlying transmission have been found to perform less well than statistical approaches to forecast dengue (30). We analyse 20 years of data from Singapore to understand the relative impact of climatic and serotype dynamics on dengue risk and to produce probabilistic forecasts from 0 - 8 weeks ahead. Our approach integrates a proxy for changes in population immunity and the epidemic potential of the dominant virus in circulation, within a statistical framework, aiming to generate accurate probabilistic predictions of dengue incidence through improved inference.

The impacts of climatic variables on dengue risk in Singapore were complex, with non-linear and delayed effects. We found increased risk of dengue at a maximum temperature of 32 °C and at intermediately wet conditions, with decreasing risk in very hot and very dry conditions. This suggests that dengue seasonality in Singapore may change as climate change leads to increasing temperatures, with fewer cases in the middle of the year. We also found non-linearly increasing dengue risk with increasing Niño 3.4 SSTAs, reflecting increased risk during El Niño conditions. We found a non-linear relationship between the time since a switch in dominant serotype and dengue transmission. In the first two years following a switch, when population immunity to the new serotype is low, we found increased dengue risk, followed by decreased dengue risk in the subsequent 4 years as immunity to the dominant serotype increases in the population. We then found evidence of increased risk at 6+ years following a serotype switch, which likely reflects the accumulation of susceptibles in the population as well as the risk associated with the growth of a non-dominant serotype before it reaches dominance, for instance growing DENV3 prevalence in 2019. Our results are in line with previous research on climate-dengue relationships in Singapore, finding that increases in temperature and precipitation increase dengue risk, as well as El Niño conditions (16,29). However, we also found evidence of non-linearity in the temperature-dengue relationship, with decreased risk at high maximum temperatures. Xu and colleagues found that absolute humidity was a better predictor for dengue incidence than other climatic variables, due to the stability of its relationship with dengue incidence during different subperiods of serotype circulation (25). In this work, we accounted jointly for the impact of serotype circulation and local weather indicators on dengue risk, and therefore were able to estimate the marginal effects of each climatic indicator without confounding from concurrent serotype dynamics. As a result of this we found stronger evidence for a role of temperature and rainfall in transmission, with little additional benefit of humidity information.

El Niño is thought to affect dengue transmission through changes in local weather conditions. We included covariates to capture both El Niño and local weather conditions within the same model structure, following the logic that the full effect of ENSO on transmission is unlikely to be fully captured by the weather covariates included in the model. For instance, ENSO may affect humidity, which was not explicitly included in the model, or temperature and precipitation metrics other than those included in the model, potentially affecting dengue season length or timing, or the spatial spread of dengue in Singapore.

We adapted our model into a proposed early-warning framework, generating accurate forecasts at operationally useful lead times (29). We used a rigorous time series cross-validation methodology to realistically evaluate model performance for early warning. This retrospective statistical validation is a necessary step in the construction of a forecasting model for early-warning. When a model is implemented operationally, forecasts may then be used for decision-making which directly impacts dengue transmission which would then complicate the evaluation of forecast accuracy. This analysis was designed in collaboration with stakeholders in the National Environment Agency of Singapore to address key questions around local dengue transmission and offers a more interpretable forecasting framework than current LASSO based forecasts (27). Future extensions of this work could include assessing whether using reported dengue cases at the time of forecast date within the model framework improves forecasting ability for operational use. Additionally, this model could be compared to other machine learning models used to forecast dengue in Singapore, or indeed combined with other approaches in an ensemble modelling framework. This model could also be used to generate probabilistic forecasts under different scenarios (e.g. comparing forecasts with or without a switch in dominant serotype or under different possible Niño 3.4 SSTAs) to understand potential outbreak risks.

Accounting for serotype dynamics within our forecasting framework helped to explain interannual variability in dengue transmission, improved dengue case forecasts at shorter lead times (up to 4 weeks ahead) and increased outbreak detection accuracy at all lead times. By comparing our full *climate and serotype* model with models only containing *climate* or *serotype* information, we were able to identify time periods where particular covariates were improving predictions. For instance, including serotype information helped to predict decreases following peaks in 2013 and 2014, as well as the peak in 2019. Contrastingly, including climate information helped to predict peak intensity and timing in 2016, during an El Niño event. It should also be noted that while our *climate and serotype* and *climate only* models perform equally well when forecasting dengue cases at longer lead times (6-8 weeks ahead), this is largely due to the flexible random effects incorporated in the modelling framework. When generating probabilistic forecasts, the covariate models estimate yearly random effects based on data available for that year up until the forecast target week. We conducted sensitivity analysis around this, running these models without a yearly random effect. In this case, the *climate only* model performs worse than the *climate and serotype* model at all forecast horizons (Supplementary information, Figures 4-5). While this demonstrates that including serotype information is helping to predict interannual variability, it also highlights the power of a flexible random effects structure to account for unmeasured variation and improve seasonal forecasts, particularly for settings without regular virus surveillance.

Despite this, there are several limitations to this study. Our *climate and serotype* model struggled to predict peak weekly cases in 2020 and 2022. This is likely due to the increase in dengue transmission in Singapore seen under SARS-CoV-2 social distancing measures, which is not accounted for in our model framework (32). We included serotype dynamics using the time since a switch in dominant serotype as a proxy for population immunity, but it would arguably be preferable to include measures of seroprevalence or estimates of the proportion susceptible directly. We were also unable to include serotype-specific dynamics due to the low number of switches in dominant serotype, even within a 17 year period of virus surveillance. This could be important to include as certain serotypes are associated with greater clinical severity and secondary infections are thought to be associated with increased severity as a result of antibody dependent enhancement (27, 28). Similarly, we only consider switches in antigenic serotype and don’t consider changes in genotype prevalence, which are also hypothesised to increase outbreak risk (35). We did not incorporate vector density into the model, which mediates the relationship between climatic conditions and dengue transmission. We did not have access to data on vector control efforts, therefore any effects of interventions would be accounted for via yearly random effects. However, it should be noted that vector control is stringently maintained within Singapore and vector density has been low for several decades. We also do not account for the early phase of pilots of Wolbachia *Aedes* suppression control strategy in Singapore with targeted releases from May 2020. This has demonstrated effectiveness in reducing mosquito populations and dengue cases, and will likely impact climate-dengue relationships and serotype dynamics in the future (36). Finally, we conducted model evaluation with forecast skill metrics and ROC analysis. In an ideal implementation scenario, forecast outputs such as outbreak warning alerts would be tied to specific control interventions or public health actions (28). This would allow for the cost of a false alarm or missed event to be calculated and enable a cost-effectiveness analysis of the early warning system as a whole.

Our analysis disentangles the role of climate and serotype dynamics in driving dengue outbreaks over a 23-year period in Singapore, capitalising on a rich dataset of epidemiological, weather station and virus surveillance data. We translate these findings into an early warning framework able to forecast dengue cases and generate outbreak alert predictions with 0-8 weeks lead time. In this study, we integrate explanatory and predictive modelling approaches with a view that understanding the key causal relationships underlying transmission allows for the construction of forecasting models that are more generalisable (37). This can also result in greater interpretability, allowing for clearer communication of forecasts to policymakers and a more intuitive understanding of how transmission may vary under future large-scale changes, such as climate change. Climate-driven early warning systems will become increasingly important adaptation measures as climate change alters the geographical range of dengue transmission and leads to greater climatic extremes. We demonstrate the additional value of viral surveillance in improving forecast accuracy, and particularly in addressing the challenge of predicting dengue outbreak years.

## Methods

### Data

In Singapore, dengue case reporting by clinicians and laboratories is mandatory. Weekly laboratory-confirmed cases from 1 January 2000 – 31 December 2022 were provided by the Ministry of Health, Singapore (14). Laboratory confirmation is performed through antigen detection of nonstructural protein 1 (NS1) or detection of viral RNA by polymerase chain reaction (PCR) in the first five days of illness, or serological detection of immunoglobulin M (IgM) after five days of illness (7). Since 2006, the National Environment Agency’s (NEA) Environmental Health Institute has serotyped a subset of dengue samples using RT-PCR as part of a virus surveillance programme (13). In our dataset, for years where serotype information is available, ∼27% of reported dengue cases are serotyped. Weekly DENV1-4 frequencies and total number of serotyped samples were provided by the Ministry of Health (14). We calculated a smoothed proportion for DENV1-4 for each week with a GAM multinomial logistic regression using *mgcv 1.8.36* (38). To smooth the serotype data, we only used data up to and including the week of interest to enable resulting models to be useful in a forward-looking early warning framework. We then use these smoothed DENV proportions to classify the dominant serotype for each week. We defined a switch event as occurring where the current dominant serotype is different from the dominant serotype in the previous week, identifying 4 switch events in our dataset. We then defined the time since a switch in dominant serotype as the number of weeks since the most recent switch event.

Midyear population size estimates were obtained from the Singapore Department of Statistics, which included both local and foreigner populations.

Weekly maximum, minimum and mean temperature (°C), absolute (g/m^3^) and relative humidity (%), and precipitation (mm) were provided by the NEA. Daily precipitation (mm) was also provided and used to calculate: the number of days without rain per week (calculated as ∑ *P_d_* = 0, where *P*_*d*_ represents rainfall on a given day); number of days with heavy rain per week (calculated as ∑ *P*_*d*_ ≥ 40); number of days with moderate to heavy rain per week (calculated as ∑ *P_d_* ≥ 20); and number of days with consecutive rainfall (i.e. a count of consecutive days where *P_d_* ≥ 1). Thresholds were chosen based on exploratory analysis of daily rainfall data and broadly align with 90th and 97.5th percentiles. Weekly Niño 3.4 SSTA was obtained from the National Oceanic and Atmospheric Administration (NOAA) (39). This index measures the El Niño Southern Oscillation (ENSO); interannual fluctuations in the oceanic and atmospheric temperature around the Pacific Ocean. The index is commonly used to define El Niño events (unusually warm) and La Niña events (unusually cool). Sea surface temperature anomalies are calculated by subtracting the observed sea surface temperatures from a historical mean, calculated for the period 1981 - 2010. El Niño and La Niña events are typically defined by a sea surface temperature anomaly of +/- 0.5°C for over 6 months.

### Model framework

We used a Bayesian hierarchical mixed-effects model to produce probabilistic predictions of weekly dengue incidence. Inference was performed using integrated nested Laplace approximation in INLA 23.04.24 (40). Weekly dengue counts (*y*_*t*_) were assumed to follow a negative binomial distribution to account for overdispersion in the data, with a mean µ_*t*_ and overdispersion parameter κ.

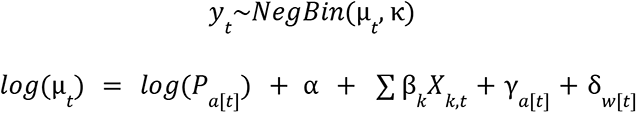

Here *log*(µ) is the linear predictor, where *log*(*P*_*a*[*t*]_) is a population offset with population per 100,000 by year and α is the model intercept . ∑ β_*k*_ 𝑋_*k,t*_ is a vector of *k* climate and/or serotype covariates, γ_*a*[*t*]_ is a yearly random effect and δ_*w*[*t*]_ is a weekly random effect. We use the weekly random effect to account for seasonality and seasonal autocorrelation, while the yearly random effect accounts for any unexplained interannual variation in the data, for example as a result of vector control efforts or COVID-19 restrictions (32). Details on model prior specifications and hyperparameters are available in the Supplementary Materials.

### Model selection

We calculated Pearson’s rank correlation index to assess correlation between variables using *corrplot 0.92* and considered *r* ≥ 0. 5 as indicative of high correlation. We also calculated the variance inflation factor to assess multicollinearity, considering *VIF* ≥ 5 as evidence of high collinearity. Based on this, we excluded relative humidity from further analysis due to high correlation with temperature variables and rainfall.

We tested all remaining variables with a 0, 4, 8, 12 and 16 week lag. For temperature, humidity and Niño variables we tested 1, 4, 8 and 12 week running averages, while for precipitation variables we tested 1, 4, 8 and 12 week running totals. Finally, we also tested non-linear formulations of temperature, precipitation and Niño 3.4 variables. We explored the best combinations of different classes of covariate (temperature, precipitation, humidity and Niño), conducting model selection in a forward stepwise manner, comparing models of increasing complexity (Supplementary information, Table 2). Covariate models were compared to a baseline model including only weekly and yearly random effects (γ_*a*[*t*]_ + δ_*w*[*t*]_). Overall 505 model formulations of climatic and serotype covariates were tested. We used various model adequacy criteria including: the widely applicable information criteria (WAIC) and deviance information criteria (DIC). WAIC and DIC are metrics which aim to maximise model fit while also penalising model complexity, with lower scores indicating a more parsimonious model.

Once we had selected the best performing climate model, we tested the inclusion of serotype variables including: dengue serotype proportions (individually and in combination, numeric variables), dengue serotype growth rates (individually and in combination, numeric variables), yearly or weekly dominant serotype (factor variable with four levels), serotype switch event (binary variable), time since serotype switch (numeric).

We quantified how much model covariates were able to explain interannual variation in dengue incidence by comparing the mean absolute value of the yearly random effects γ_*a*[*t*]_ between covariate models and the baseline model. We calculated the proportion of interannual variation explained by the model as:

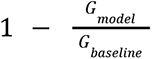

where 𝐺 = 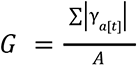 and A is the total number of years in the dataset.

### Model evaluation

We evaluated the performance of four candidate models: a *climate and serotype* model; a *climate only* model; a *serotype only* model; a *seasonal baseline* model. We conducted model evaluation using time series cross-validation methodology to produce posterior predictions, and comparing observed and predicted outcomes to evaluate predictive performance. This is an appropriate cross-validation design when conducting statistical validation of forecasting models, as it preserves the time order of the underlying data, i.e. only data prior to an observation occurring is used to generate the prediction. To do this, we refit the model for each week in the dataset from 2009-2023 using an expanding window approach (31). Data for the first eight years was used solely for training. Then, for each target week in the dataset, we trained the model on data until week *t* − 1 and then simulated a posterior predictive distribution for dengue cases in week *t*, using climatic data up until time *t*. Serotype covariates are constructed only using data until *t* - 1 for each time point as serotype frequencies are linked to dengue case counts. The posterior predictive distribution was simulated using 1000 samples from the posterior distribution of model parameters and hyperparameters. Note that all our final models (except the *seasonal baseline*) included a yearly random effect, γ_*a*[*t*]_, which is estimated using only using data available until week for the year of the target week.

We also calculated posterior predictive probabilities of exceeding the outbreak threshold. This both evaluates the model’s ability to distinguish between outbreak and non-outbreak periods, and provides an operationally useful model output for decision makers. For instance, it may be easier to tie specific response measures or public health decisions to a probability of exceeding an operationally meaningful threshold than to the full probabilistic forecast. Here, we calculated outbreak thresholds for each week based on the seasonal moving 75th percentile of cases (Supplementary information, Figure 1). For each week *t*, we then calculated the proportion of posterior predictive samples that were greater than the threshold value.

Forecasts were scored using the *scoringutils 1.2.1* package (41). We aimed to evaluate both how successful models were at predicting dengue case incidence and detecting outbreak weeks. Dengue case forecasts were scored using the continuous ranked probability score (CRPS); this is a proper scoring rule which can be considered a generalisation of mean absolute error that takes into account the entire predictive distribution (42). Smaller CRPS values indicate a better forecast, and the metric penalises both under- and over-prediction. Sharper forecasts (where predictions are concentrated in a narrower range) will also score better. We then calculated the continuous ranked probability skill score (CRPSS), which is calculated as 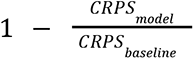 and measures the percentage improvement of the considered model over a baseline model. A value of 1 indicates that the model has perfect skill compared to the baseline, 0 indicates the model is equivalent to the baseline and a negative value indicates that the model is worse than the baseline.

We also assessed model calibration by calculating interval coverage at the 50% and 95% levels. Interval coverage measures the proportion of observed values falling in a given prediction interval range. For a given prediction interval, a perfectly calibrated model would have interval coverage equal to the nominal prediction interval (that is, 95% of observations falling within the 95% prediction interval). We also calculated bias 𝐵, measuring a model’s tendency to over or under-predict. This was calculated for a data point *y*_t_ such that:

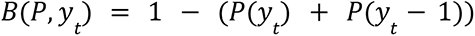

Where *P*(*y*_t_) is the predicted probability mass for all outcomes smaller or equal to *y*_t_.

To score the models’ ability to forecast future outbreaks we calculated the Brier score comparing the posterior predictive probability of exceeding the outbreak threshold with observed outbreak weeks. The Brier score is a proper scoring rule for binary outcomes computed as the mean square error between the probabilistic prediction and the true outcome, where smaller values indicate better forecasts (43). Finally, we used receiver operating characteristic (ROC) analysis to determine the optimum threshold for issuing outbreak alerts, balancing hit rate with false alarm rate, using the *pROC package 1.18.4* (44,45). Here, hit rate (or sensitivity) is defined as the proportion of events (outbreak weeks) that were correctly predicted. False alarm rate (or 1 - specificity) is defined as the proportion of weeks without an outbreak where an outbreak was predicted to occur. We generated ROC curves for each model, which show hit rate against false alarm rate at different outbreak alert thresholds and calculated the area under the ROC curve (AUC). The AUC is a measure of model performance in classifying outbreak and non-outbreak weeks, with higher values indicating a better classification (46). We selected outbreak alert thresholds by choosing the point closest to the top left of the ROC plot (representing perfect sensitivity or specificity).

### Adapting the model for early warning

To adapt our model framework for use in an early warning scenario, we produced and evaluated dengue case forecasts at 2, 4, 6 and 8 week ahead forecast horizons. To do this, we used the best approximation of each covariate used in the final models available at the forecast horizon being considered. For instance, as in our final models we use a 12 week running average of maximum temperature (°C), to produce forecasts, at a 4 week ahead time horizon we instead use an 8 week running average of maximum temperature (°C), with a four week lag. As the rainfall covariate we include is a 12 week total of days without rain, to approximate this for a 4 week ahead time horizon we use an 8 week total of days without rain, scaled up by a factor of 1.5. Full details of the variables used for prediction at each time horizon are available in Supplementary information, Table 5.

When conducting model evaluation for week *t* and forecast horizon *h*, we used the same expanding window time series cross-validation approach described earlier. We trained the model using the final selected covariates on data available until week *t* − *h*. Unlike the model evaluation design described earlier, we then predicted dengue incidence in week *t* using lagged covariates available at time *t* − *h*. For example, considering only the temperature covariate for simplicity, when predicting dengue cases with a 4 week ahead time horizon, we fit the model up until week *t* − *h* using a 12 week running average temperature to estimate model parameters. Then, using these estimated model parameters, we predict dengue cases at week *t* by inputting 8 week running average temperature with a 4 week lag (alongside other lagged covariates). A schematic showing the cross-validation design for different forecast horizons is shown in Supplementary Figure 3. This approach allows us to preserve the key relationships between climate and serotype covariates, and dengue cases that we estimate in full model fitting and then use the best climate data available at different lead times to generate forecasts for early warning.

## Code availability

All code and data used for this analysis are available at: https://github.com/EmilieFinch/dengue-singapore

## Author Contributions

Conceptualisation: EF, RL, SS, LCN. Methodology: EF, RL, AJK. Data curation: SS, LCN, EF. Investigation: EF, RL. Supervision: RL, AJK. Data analysis: EF. Figures: EF. Writing - original draft: EF, RL, AJK. Writing - review and editing: all authors.

