## Supplementary Material for "Climate variation and serotype competition drive dengue outbreak dynamics in Singapore"

**Model priors and hyperparameter formulations**

The weekly random effect $\delta_{w[t]}$ was assigned a random walk 2 prior distribution (second order difference prior distribution) where we assume that second order increments follow a Gaussian distribution with zero mean and precision $\tau$. This was defined to be cyclic (so the first week of the year is dependent on the last two weeks of the previous year). We included independent and identically distributed (iid) random effects for each year $\gamma_{a[t]}$. Non-linear covariates were modelled by setting a random walk 2 prior on the coefficients of the covariates. We used penalized complexity priors (PC priors) for precision $\tau$, with hyperparameters $\sigma_0 = 0.5$ and $\alpha = 0.01$ for all non-linear covariates and random effects in the model. The PC prior is defined on the standard deviation $\sigma = \tau^{-1/2}$ such that $P(\sigma > \sigma_0) = \alpha$. these penalize departure from $\sigma = 0$. PC priors follow the principle of parsimony, favouring a base model (where $\sigma = 0$) unless evidence is provided against it (1).

**Table 1: Covariates tested in model selection**

| Variable name | Covariate class | Variable type |
|---|---|---|
| Minimum temperature °C | Temperature | Numeric |
| Mean temperature °C | Temperature | Numeric |
| Maximum temperature °C | Temperature | Numeric |
| Absolute humidity $g/m^3$ | Humidity | Numeric |
| Relative humidity % | Humidity | Numeric |
| Total precipitation (mm) | Rainfall | Numeric |
| Number of days without rain | Rainfall | Numeric or categorical |
| Number of days with heavy rain | Rainfall | Numeric or categorical |
| Number of days with moderate or heavy rain | Rainfall | Numeric or categorical |
| Number of days with consecutive rainfall | Rainfall | Numeric or categorical |
| Niño 3.4 sea surface temperature anomalies (SSTA) | ENSO | Numeric |
| Serotype proportions | Serotype | Numeric |
| Serotype growth rates | Serotype | Numeric |
| Dominant serotype | Serotype | Factor |
| Switch in dominant serotype | Serotype | Binary |
| Time since switch in dominant serotype | Serotype | Numeric |

**Table 2: Details of model selection**

We conducted forwards stepwise selection, grouping climatic indicators into classes of covariate including: temperature, rainfall, humidity and Niño 3.4. At each step of model selection, the best performing variable was carried forwards and tested against all variables in remaining climate classes. Note that at the third stage of model selection of model selection the 12 week total days without rain performed similarly well to 12 week total precipitation (mm); we selected the former due to the high influence of outlier precipitation values on the estimated effect size. Finally, we did not include absolute humidity as there was evidence for no effect on dengue incidence and only marginal improvements in model adequacy criteria.

| Step | Variable | WAIC | DIC | Rsq |
|---|---|---|---|---|
| 1 | Maximum temperature (12 week running average, non-linear, 0 lag) | 12830.96 | 12829.96 | 0.067 |
| 2 | Maximum temperature (12 week rolling average, 0 lag) + Niño 3.4 (12 week rolling average, non-linear, 1 month lag) | 12786.84 | 12785.10 | 0.103 |
| 3 | Maximum temperature (12 week rolling average, 0 lag) + Niño 3.4 (12 week rolling average, non-linear, 1 month lag) + 12 week total days without rain (0 lag) | 12756.24 | 12754.75 | 0.139 |
| 4 | Maximum temperature (12 week rolling average, non-lineaer, 0 lag) + Niño 3.4 (12 week rolling average, non-linear, 1 month lag) + 12 week total days without rain (non-linear, 0 lag) + absolute humidity (4 month lag) | 12748.57 | 12746.83 | 0.146 |
| Adding serotype | Maximum temperature (12 week rolling average, non-linear, 0 lag) + Niño 3.4 (12 week rolling average, non-linear, 1 month lag) + 12 week total days without rain (non-linear, 0 lag) + time since switch in dominant serotype (non-linear) | 12498.13 | 12498.27 | 0.331 |

### Table 3: Details of selected models

Full model formulae for full *climate and serotype* model and other models compared in the main text.

| Final models | Formula |
|---|---|
| *Climate and serotype* | Time since switch in dominant serotype (non-linear) + maximum temperature °C (12 week average, non-linear) + days without rain (12 week total, non-linear) + Niño 3.4 SSTA (12 week average with a 4 week lag) + $\gamma_{a[t]} + \delta_{w[t]}$ |
| *Climate only* | Maximum temperature °C (12 week average, non-linear) + days without rain (12 week total, non-linear) + Niño 3.4 SSTA (12 week average with a 4 week lag) + $\gamma_{a[t]} + \delta_{w[t]}$ |
| *Serotype only* | Time since switch in dominant serotype (non-linear) + $\gamma_{a[t]} + \delta_{w[t]}$ |
| *Seasonal baseline* | $\delta_{w[t]}$ |

### Table 4: Forecast metrics over different forecast horizons

Forecast skill metrics for each candidate model at forecast horizons from 0-8 weeks. Metrics include: CRPS (lower scores are better) CRPSS (higher scores are better), Brier score (lower scores are better), AUC (higher scores are better), false alarm rate (lower scores are better) and hit rate (higher scores are better). The trigger threshold which maximises the AUC for each model is also shown. The best score for each forecast metric and horizon is shown in bold.

| Horizon | Model | CRPS | CRPSS | Brier score | AUC | False alarm (%) | Hit rate (%) | Trigger threshold (%) |
|---|---|---|---|---|---|---|---|---|
| 0 | *Climate and serotype* | **50** | **59.5** | **0.0544** | **98.4 (95% CI: 97.69-99.03)** | **2.08** | **91.6** | 71.4 |
| 0 | *Serotype only* | 63.2 | 48.9 | 0.062 | 97.8 (95% CI: 97-98.65) | 2.54 | 90.7 | 65.4 |
| 0 | *Climate only* | 57.5 | 53.5 | 0.0655 | 97.8 (95% CI: 96.95-98.6) | 3.92 | 93.4 | 54.7 |
| 0 | *Seasonal baseline* | 124 | 0 | 0.232 | 73.4 (95% CI: 69.8-77.09) | 11.8 | 65.2 | 37.8 |
| 2 | *Climate and serotype* | **56.4** | **54.9** | **0.0623** | **97.8 (95% CI: 96.96-98.58)** | 3.46 | 91 | 64 |
| 2 | *Serotype only* | 70 | 43.9 | 0.0713 | 97 (95% CI: 95.99-98.03) | **2.77** | 88.9 | 65.6 |
| 2 | *Climate only* | 62.3 | 50.1 | 0.0722 | 97 (95% CI: 96.05-98.03) | 4.38 | **92.5** | 49.6 |
| 2 | *Seasonal baseline* | 125 | 0 | 0.235 | 71 (95% CI: 67.22-74.79) | 14.6 | 66.7 | 37 |

| | | | | | | | | |
|---|---|---|---|---|---|---|---|---|
| 4 | *Climate and serotype* | **67.9** | **46.3** | **0.0773** | **96.5 (95% CI: 95.44-97.62)** | 5.54 | **90.4** | 49.2 |
| 4 | *Serotype only* | 82.4 | 34.9 | 0.0894 | 94.9 (95% CI: 93.45-96.4) | 5.19 | 87.1 | 50.6 |
| 4 | *Climate only* | 71.7 | 43.4 | 0.0863 | 95.4 (95% CI: 94.11-96.78) | **5.07** | 89.5 | 49.2 |
| 4 | *Seasonal baseline* | 127 | 0 | 0.24 | 68 (95% CI: 64.11-71.94) | 15.6 | 64.3 | 37 |
| 6 | *Climate and serotype* | 86.5 | 32.4 | **0.0889** | **95.4 (95% CI: 94.1-96.74)** | 6.23 | **89.5** | 45.4 |
| 6 | *Serotype only* | 104 | 18.8 | 0.105 | 93.1 (95% CI: 91.31-94.86) | 5.77 | 83.2 | 54.6 |
| 6 | *Climate only* | **86.2** | **32.7** | 0.0982 | 94 (95% CI: 92.37-95.57) | **4.38** | 83.5 | 52.5 |
| 6 | *Seasonal baseline* | 128 | 0 | 0.243 | 65.3 (95% CI: 61.35-69.35) | 17.1 | 64.6 | 36.2 |
| 8 | *Climate and serotype* | 86.5 | 32.4 | **0.098** | **94.2 (95% CI: 92.65-95.75)** | 6.92 | **90.7** | 41.7 |
| 8 | *Serotype only* | 104 | 18.8 | 0.123 | 90.9 (95% CI: 88.79-92.98) | 7.38 | 81.4 | 51.5 |
| 8 | *Climate only* | **86.2** | **32.7** | 0.115 | 92.1 (95% CI: 90.24-93.92) | **6** | 80.5 | 51.6 |
| 8 | *Seasonal baseline* | 128 | 0 | 0.245 | 64.3 (95% CI: 60.27-68.33) | 14.2 | 56.5 | 37.8 |

CRPS: continuous ranked probability score; CRPSS: continuous ranked probability skill score; AUC: area under the curve; CI: confidence interval

**Table 5: Variables used for prediction at each time horizon**

| Forecast horizon | Temperature | Precipitation | ENSO | Serotype |
|---|---|---|---|---|
| 0 | 12 week average maximum temperature °C, 0 week lag | 12 week total days with no rain, 0 week lag | 12 week rolling average Niño 3.4 SSTA, 4 week lag | Time since serotype switch, 0 week lag |
| 2 | 10 week average maximum temperature °C, 2 week lag | 10 week total days with no rain, 2 week lag, scaled up by 1.2 | 12 week rolling average Niño 3.4 SSTA, 4 week lag | Time since serotype switch, 2 week lag + 2 |
| 4 | 8 week average maximum temperature °C, 4 week lag | 8 week total days with no rain, 4 week lag, scaled up by 1.5 | 12 week rolling average Niño 3.4 SSTA, 4 week lag | Time since serotype switch, 4 week lag + 4 |
| 6 | 6 week average maximum temperature °C, 6 week lag | 6 week total days with no rain, 6 week lag, scaled up by 2 | 10 week rolling average Niño 3.4 SSTA, 6 week lag | Time since serotype switch, 6 week lag + 6 |
| 8 | 4 week average maximum temperature °C, 8 week lag | 4 week total days with no rain, 8 week lag, scaled up by 3 | 8 week rolling average Niño 3.4 SSTA, 8 week lag | Time since serotype switch, 8 week lag + 8 |

**Figure 1: Dengue cases and outbreak threshold**

Figure showing weekly reported dengue cases from 1 January 2000 – 31 December 2022 (grey bars) and seasonal moving 75th percentile outbreak threshold (green line) and the endemic channel threshold used operationally by the NEA (brown line). For a given month and year, we defined a seasonal moving 75th percentile outbreak threshold using the 75th percentile of weekly cases in that month using all years up to, but not including, the current year. The NEA uses an endemic channel threshold which is calculated as two standard deviations in excess of mean cases over the past 5 years, with outliers removed. Outliers are defined as any weekly cases greater than the threshold for that year.

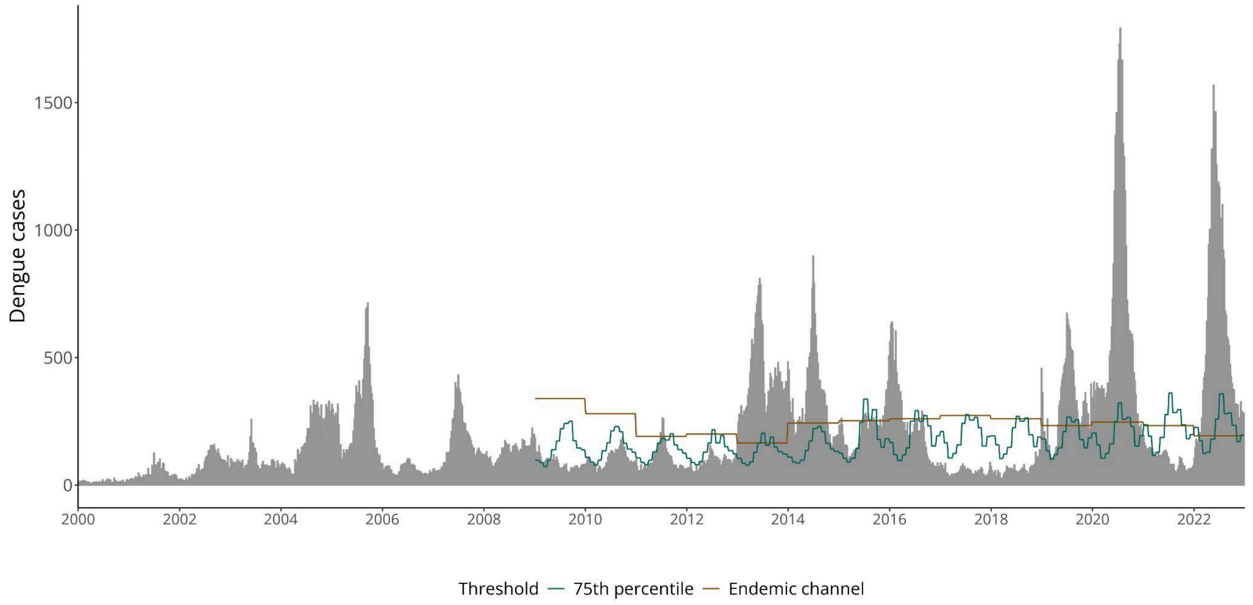

**Figure 2: Dengue forecasts for early warning at 2-8 week forecast horizons**

Figure showing time series of cross-validated posterior predictions of dengue cases for each model from 2009 – 2022 at 2 - 8 week forecast horizons. Columns for 4 weeks ahead and 8 weeks ahead are also shown in Figure 5 of the main text. Coloured lines show the median posterior prediction of weekly dengue cases, shaded areas show the 95% credible interval and the dark grey line shows the data. From top to bottom the figure shows: predictions for the final selected '*Climate and serotype*' model weekly and yearly random effects $\gamma_{a[t]} + \delta_{w[t]}$ in purple; predictions for a '*Climate only*' model with random effects in pink; predictions for a '*Serotype only*' model with random effects in green; and predictions from a '*Seasonal baseline*' model with only weekly random effects $\delta_{w[t]}$ in orange. From left to right each column shows forecasts at 2, 4, 6 and 8 weeks ahead respectively.

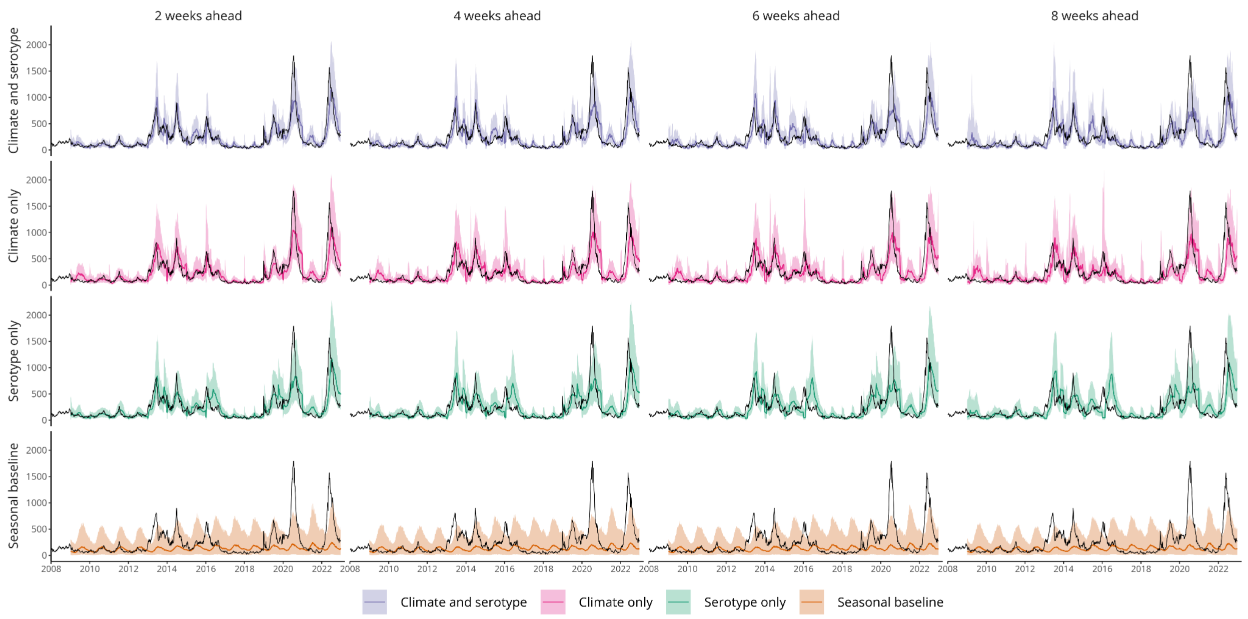

**Figure 3: Schematic of time series cross validation design to generate forecasts**

Schematic showing how $h$ week ahead forecasts are generated using time series cross-validation. Blue blocks represent the training data, orange blocks time points to be forecasted and grey points represent data not included in the forecast generation. Columns show the training/testing design for 2-step ahead and 3-step ahead forecasts respectively, while rows show different example municipalities. We use an expanding window approach where, for each forecast horizon $h$, we train the model on data available until week $t$ - $h$. We then predict dengue incidence in week $t$, using lagged covariates. When conducting model evaluation for week $t$ and forecast horizon $h$, we used the same expanding window time series cross-validation approach (Methods). We trained the model using the final selected covariates on data available until week $t - h$. We then predicted dengue incidence in week $t$ using the best approximation of each climatic covariate. For example, considering only the temperature covariate for simplicity, when predicting dengue cases with a 4 week ahead time horizon, we fit the model up until week $t - h$ using a 12 week running average temperature to estimate model parameters. Then, using these estimated model parameters, we predict dengue cases at week $t$ by inputting 8 week running average temperature with a 4 week lag (alongside other lagged covariates). This allows us to preserve the key relationships between climate and serotype covariates, and dengue cases that we estimate in full model fitting and then use the best climate data available at different lead times to generate forecasts for early warning.

Note that this differs from the design used for model evaluation (shown in Figure 3 of the main text), where data until $t - 1$ is used to train the model. Climatic data up to time $t$ is then used to generate predictions for dengue incidence at time $t$. Serotype covariates are constructed using data until $t - 1$ as serotype frequencies are dependent on case counts.

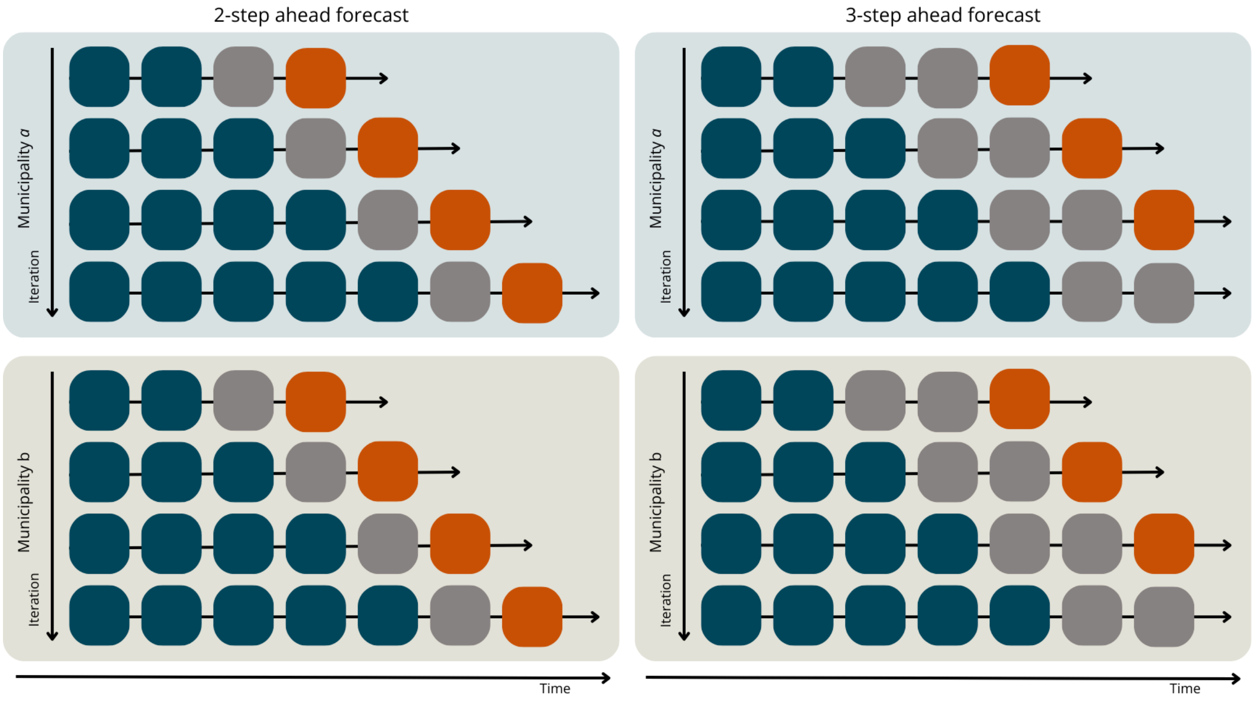

**Figure 4**: **Dengue forecasts for early warning at 2-8 week forecast horizons showing covariate models without yearly random effects**

Figure showing time series of cross-validated posterior predictions of dengue cases for each model from 2009 – 2022 at 2 - 8 week forecast horizons. Here, unlike Figure 5 in the main text, the *Climate and serotype*, *Climate-only* and *Serotype-only* models do not include a yearly random effect $\gamma_{a[t]}$. Coloured lines show the median posterior prediction of weekly dengue cases, shaded areas show the 95% credible interval and the dark grey line shows the data. From top to bottom the figure shows: predictions for the final selected '*Climate and serotype*' model with weekly random effects $\delta_{w[t]}$ in purple; predictions for a '*Climate only*' model with weekly random effects in pink; predictions for a '*Serotype only*' model with weekly random effects in green; and predictions from a '*Seasonal baseline*' model with only weekly random effects in orange. From left to right each column shows forecasts at 2, 4, 6 and 8 weeks ahead respectively.

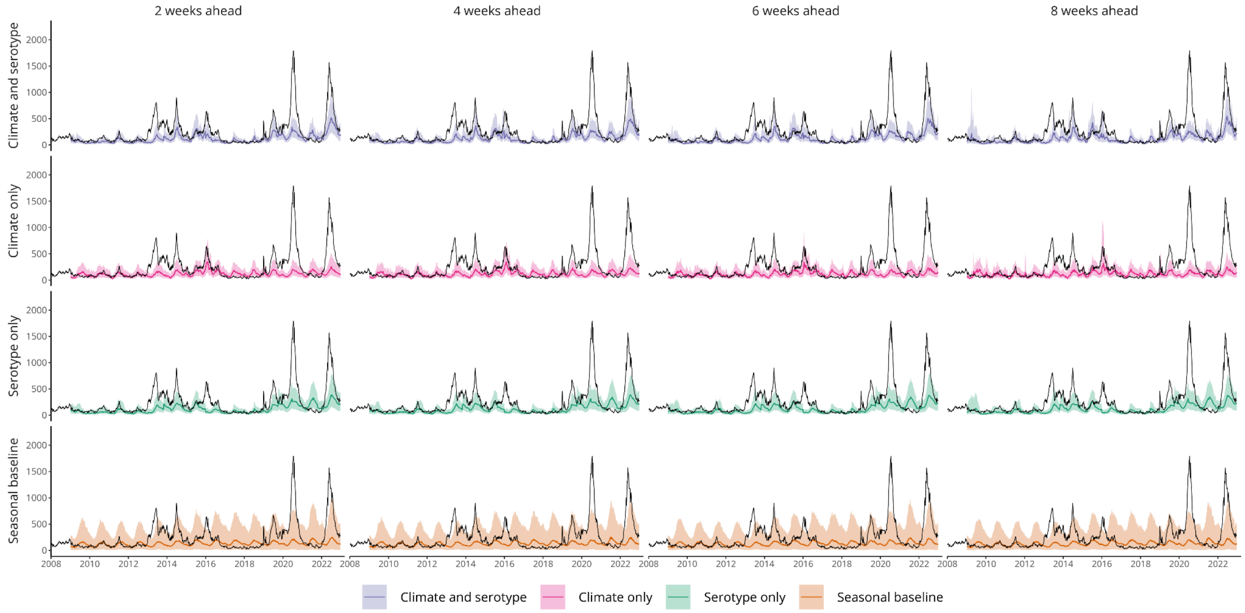

**Figure 5: Predictive performance over different forecast horizons for covariate models without yearly random effects.** Figure showing forecast metrics for each model across all forecast horizons from 2009 – 2022. Unlike Figure 6 in the main text, here covariate models do not include a yearly random effect $\gamma_{a[t]}$. Figure showing forecast metrics for each model across all forecast horizons from 2009 – 2022. From top left to bottom right these show: interval coverage %, bias; CRPS (continuous ranked probability score), CRPSS (continuous ranked probability skill score, %), Brier score, AUC (area under the curve, %), hit rate (%) and false alarm rate (%). Interval coverage shows the percentage of observations falling inside a given prediction interval. A perfectly calibrated forecast would have coverage equal to the nominal prediction interval; that is, 95% coverage equal to 95% and 50% coverage equal to 50%, indicated by dashed horizontal lines. Bias measures the relative tendency of the model to over- or under-predict, and is bounded between -1 and 1, with 0 indicating unbiased forecasts. The CRPS can take values between 0 and infinity, with smaller values indicating better performance. The CRPSS indicates the relative improvement of each covariate model over the *seasonal baseline* model and can take values from 0%, indicating that the model performs the same as the baseline, and 100%, indicating perfect forecasting skill. The Brier score can take values from 0 – 1, with smaller values indicating better performance. The AUC can take values from 0-100% with 100% indicating perfect classification. Hit rate and false alarm rate also take values from 0 – 100% with higher and lower values indicating better performance respectively.

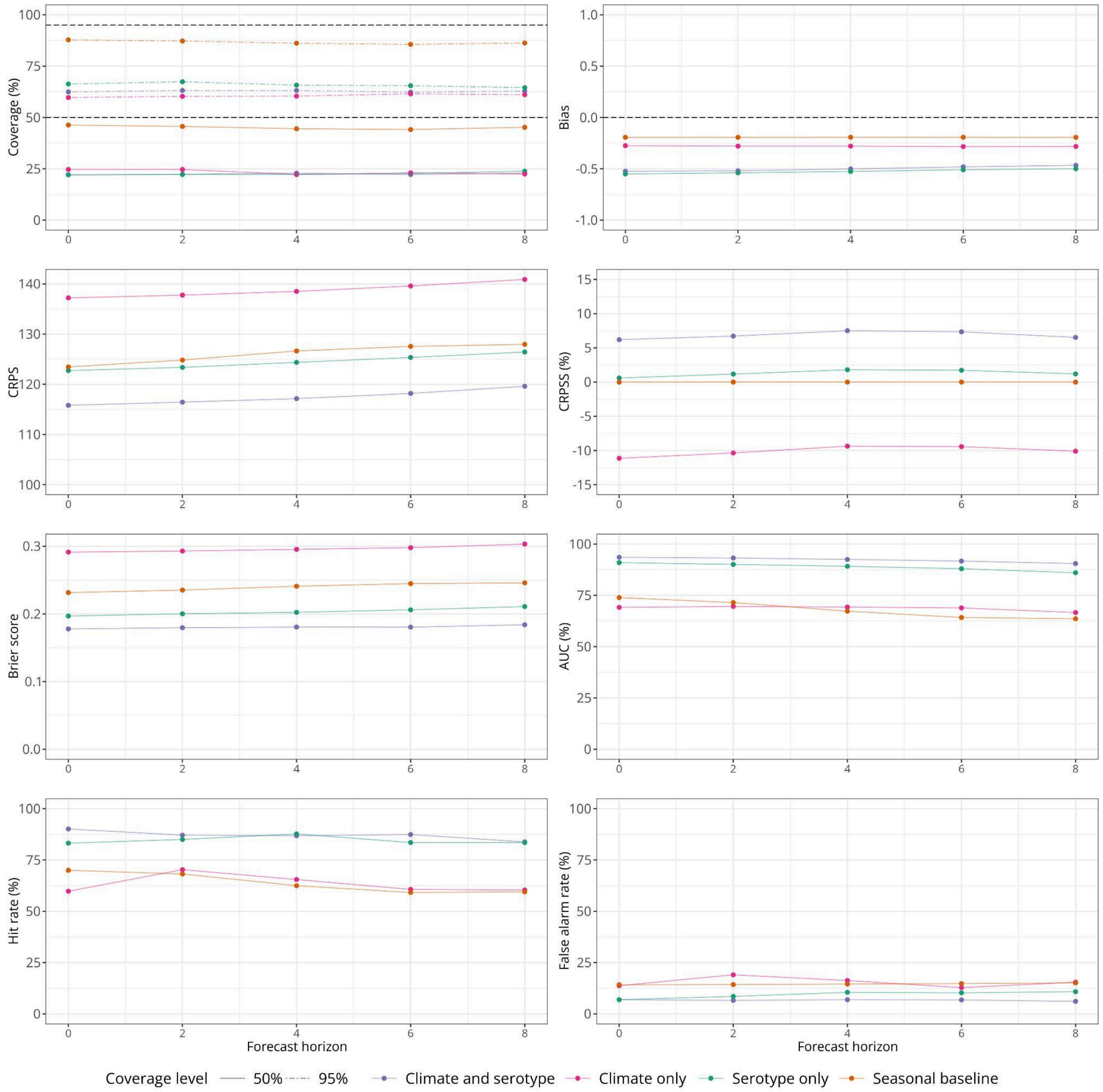
